# Staff testing in care homes for older people: policy implications for early stages of future pandemic responses

**DOI:** 10.1101/2025.06.09.25329311

**Authors:** Lily O’Brien, Oliver Stirrup, Catherine Henderson, James Blackstone, Natalie Adams, Borscha Azmi, Lavinia Bertini, Jackie Cassell, Dorina Cadar, Andrew Copas, Christopher Finn McQuaid, Paul Flowers, Laura Shallcross, Lara Goscé

## Abstract

**Background:** Care home residents are at high risk of severe outcomes following respiratory infection due to age, co-morbidities, and close contact with staff and other residents. Frequent staff testing could potentially reduce respiratory infection cases in residents, but evidence is limited. This study uses historical COVID-19 data in England to assess the impact of varying staff testing rates under different transmission scenarios to inform response during future pandemics.

**Methods:** We developed a compartmental model of SARS-CoV-2 transmission in England, with three population strata: general population, care home staff, and residents. The model was calibrated using prevalence data from January 2021 to March 2022 and testing rates from the VIVALDI Study (ISRCTN14447421). We conducted a scenario analysis projecting resident cases and deaths over 12 months under varying staff testing frequencies (monthly, twice-monthly, weekly, twice-weekly, daily) assuming a new dominant strain. We also explored the impact of testing when combined with a hypothetical low-cost highly-effective public health and social measures.

**Findings:** Staff testing alone has little impact on reducing cases and deaths in the resident population. Daily testing could avert only 3.8% (95%UI: 3.1-4.4%) cases and 3.5% (95%UI: 2.3-4.4%) deaths compared to a baseline testing of less than one test per month. When combined with public health and social measures, however, the effect is large. Daily staff testing, combined with public health and social measures, can reduce resident cases by 54% (95%UI: 50-58%) and deaths by 50% (95%UI: 26-59%). Additionally, if testing frequency is reduced but there is still a public health and social measure, the effect size decreases, however there are potential cost savings (£0.7 to £3.4 million).

**Interpretation:** Increasing staff testing alone is insufficient to significantly reduce SARS-CoV-2 cases and deaths in care home residents. However, combining testing with some form of a public health and social measure aimed at reducing transmission among residents, is epidemiologically effective and cost-effective in most scenarios and possibly cost saving.

## Introduction

Over a quarter of a million individuals aged 65 and above live in care homes in England and Wales [1] with many affected by comorbidities that increase their risk of poor outcomes. Care homes face high risks of disease outbreaks due to several factors, including the challenges of social distancing in shared living spaces and communal areas, and frequent contact required between staff and residents for daily care. Between March 2020 and May 2021, 24.3% of deaths of care homes residents had been attributed to COVID-19 in England and Wales, with 42,341 total deaths involving COVID-19 being registered [2]. It is essential that care homes can function safely and efficiently during future pandemics to protect vulnerable residents and care home staff. COVID-19 vaccinations were made available to high-risk groups, including care home residents and staff, from December 2020 [2], providing protection against the virus. However, before the introduction of vaccines in England, care homes faced high cases and deaths amongst residents. This illustrates the importance of having robust policies and procedures in place to minimise disruptions to the care home sector; protecting vulnerable populations especially at the beginning of a new respiratory pandemic when pharmaceutical interventions may not be available yet.

While several studies have examined intervention strategies in care homes [3–5] the findings vary regarding the most effective approaches to minimising resident cases and deaths, with some studies recommending asymptomatic staff testing every 3 days [3] and others every 7 to 10 days [4]. Additionally, some research highlights the importance of pharmaceutical interventions such as vaccination, illustrating its increased efficacy over staff testing [6] and others emphasise the importance of other public health and social measures such as isolating positive cases [4], personal protective equipment (PPE), improved ventilation and contact tracing. It is crucial to determine the best strategies for outbreak control, while also understanding the associated economic costs and quality of life of residents, to guide policy decisions in future pandemics.

In this study, we aim to assess possible testing policies to be adopted during future pandemics. To do so, we used COVID-19 as a case study and developed a compartmental model simulating SARS-CoV-2 transmission in care homes in England. While we modelled COVID-19, due to it being the latest pandemic and having available data, the model is adaptable to similar respiratory infections and could be highly informative in future pandemics, including avian influenza (in case of human-to-human transmission) or emerging mpox variants. The aim of our study is to provide evidence for effective testing protocols that can enable early detection and infection control in vulnerable populations such as care homes. We compared a baseline scenario with three intervention strategies to determine the optimal frequency of staff testing and assess its effectiveness when combined with a public health and social measure. Additionally, we took associated testing and hospitalisation costs into account and estimated the economic impact of these interventions in the form of incremental costs per case averted.

## Methods

### Model Overview

We developed a compartmental model to simulate the transmission of SARS-CoV-2 in England, stratifying the population into three distinct groups: the general population, care home staff, and care home residents. The general population encompasses all individuals in England who are not classified as care home staff or residents, including all age groups. Care home staff are defined as individuals aged 18 and over who work in care homes in direct contact with residents, while care home residents are aged 65 and above. The model considers a cohort comprising 10,000 senior care home residents and estimated 13,350 staff members by calculating the ratio of staff and occupied beds (∼1:1.3) from the VIVALDI Study (ISRCTN14447421) [7].

The Vivaldi-CT trial which informed this analysis began in January 2023 and was developed in close collaboration with care providers. Engagement events with a diverse range of care home staff and sector representatives helped shape the trial intervention “Test to Care”, supporting twice-weekly LFD testing for COVID-19 in care homes. Discussions with care providers addressed the acceptability and uptake of testing and acted as a platform to raise concerns about potential implications for staff members and organisations involved in the trial. Feedback informed the testing protocol. However, due to lower than anticipated COVID-19 case numbers, the trial was stopped early. Here, we use mathematical modelling to simulate the original trial design and assess how it might perform in the early stages of a future pandemic.

A critical assumption of our model is that care homes operate at full capacity, with new residents replacing those who have died. Because we want our model to be informative of the first stages of a new pandemic, we assume that residents are unvaccinated and lack long-term immunity from previous infections. Additionally, to reflect the restriction in visitations allowed at the beginning of the COVID-19 pandemic, we assume that while staff interact with the general population, residents, and other staff, residents only interact with staff and fellow residents. Consequently, the primary routes of SARS-CoV-2 ingress into the care home setting are via staff or new residents, excluding visitors or contractors. This model thus reflects a pandemic scenario restricted to internal transmission within the care home environment. We based this assumption on observations from the COVID-19 pandemic, during which contact between care homes and the outside world was significantly restricted to mitigate transmission risks. We acknowledge however that these restrictions may differ for future pandemics, which could affect the applicability of our findings.

The model, which employs a susceptible-exposed-infectious-recovered (SEIR) framework, was coded using R (v4.3.3) and calibration was performed through the History Matching and Emulation (hmer) package [8–9]which uses Bayes Linear emulation and history matching, The model was calibrated to observed COVID-19 prevalence rates in the general population [10]and care home residents [7] between January 2021 and March 2022. Full parametrisation, as well as model schematic, equations, and calibration, can be found in the supplementary document.

Following calibration, we split our model into a baseline scenario and different intervention scenarios, with testing rates and the implementation of an additional public health and social measure differing between them. Due to new strains differing in terms of infectiousness and mortality [11], the objective was to consider a new strain to inform policy making for future pandemics. Therefore, we modelled a new hypothetical strain that is both more infectious and more deadly. We then estimated resident cases and death for the following 12 months.

### Intervention scenarios

We evaluated three intervention scenarios against a baseline, as shown in Table 1. The baseline scenario assumes that staff members are tested less than once a month (∼0.6), in order to represent a hypothetical baseline situation where only symptomatic individuals are required to test. Intervention 1 scenarios, instead, consider the following five testing regimes: (a) monthly, (b) twice a month, (c) once a week, (d) twice a week and (e) daily testing of all staff. In all scenarios, lateral flow tests (LFT) for staff were assumed to have perfect sensitivity, meaning any positive result was treated as a true positive, with staff immediately self-isolating until recovery. The model was designed to study a pandemic period, so we also modelled two additional intervention scenarios that consider a hypothetical continuous public health and social measure aimed at reducing infection transmission among residents. Intervention 2 considers baseline testing plus a public health and social measure, while intervention 3 combines intervention 1 (the five testing regimens) and intervention 2 (the public health and social measure). To estimate the maximum possible impact of staff testing, we assumed that this hypothetical public health and social measure leads to no infection transmission among residents, whilst transmission between staff and residents is possible. All interventions are described in Table 1.

**Table 1:** Intervention scenarios and description.

| **Intervention**<br>**scenario** | **Description** |
| --- | --- |
| Baseline | Staff are tested less than once a month (~0.6 tests per month per staff member) |
| Intervention 1 | Staff are tested either (a) once a month, (b) twice a month, (c) once a week, (d) twice a week or (e) daily |
| Intervention 2 | Baseline testing with a public health and social measure |
| Intervention 3 | Staff are tested either (a) once a month, (b) twice a month, (c) once a week, (d) twice a week or (e) daily, in conjunction with a public health and social measure |

For every intervention, and testing frequency within the intervention, we calculated numbers of resident cases and deaths observed and compared them to the baseline scenario estimating numbers of cases and deaths averted.

### Health economic analysis

We associated testing costs and hospitalisation costs with the epidemiological modelling results. No costs are accounted for the public health and social measure. Testing and hospitalisation costs were accounted for in each intervention scenario (including baseline) and were associated to the modelling output ‘cases averted’, which differed depending on the testing strategy and the inclusion of a hypothetical public health and social measure. Testing costs included LFT costs, staff sick pay, and agency backfill. Staff sick pay and agency backfill costs were sourced from audit data provided by the UCL Comprehensive Clinical Trials Unit (CCTU) collected during the VIVALDI-CT [12] factoring in the average duration of staff absences. The unit costs for LFTs were obtained from the United Kingdom Health Security Agency (UKHSA) [13]. LFT costs were multiplied by the total number of care home staff assumed in the model over a 12-month period. As each scenario required different testing frequencies, the number of LFTs performed varied by the intervention (e.g. in the case of monthly testing, 13,350 tests each month over a 12-month period are assumed to be performed). Staff sick pay and agency backfill costs were calculated based on the number of staff self-isolating over the 12 months, which also differed across intervention scenarios. The average cost per staff member (averaging over mean costs per home) was *per episode* following a positive test. Only shifts that were reimbursed by UKHSA were counted. This was a sample of 24 care homes representing 180 staff members with COVID-related sickness. These costs—sick pay, agency backfill, and LFTs— were then combined to determine the total cost per intervention. All costs are reported in 2022-23 pounds (£).

Hospitalisation costs were calculated for each intervention and were divided into critical care and non-critical care costs. Resident hospitalisations were estimated from our model output, by multiplying the number of individuals who become infected with SARS-CoV-2 for each intervention, by the proportion of cases requiring either critical or non-critical care. As the aim of this analysis is to estimate the cost-effectiveness of the aforementioned testing interventions during the first stages of a new pandemic, the hospitalisation rates were based upon a report from Ferguson et al. (2020) [14] who estimated the severity of COVID-19 cases by age-group during the first wave of the pandemic in England (however we also acknowledge that in reality some residents requiring hospitalisation will not necessarily access these facilities). Critical and non-critical care costs were obtained from the National Institute for Health and Care Excellence (2023)[15]. All cost data is reported in the technical appendix. Due to the theoretical nature of interventions 2 and 3, we do not consider public health and social measure associated costs, which may include costs such as increased cleaning, re-rostering of staff, provisions of face masks, etc. and thus could not be included in this analysis.

Following this we performed a cost-effectiveness analysis by estimating the incremental cost of the interventions (compared to baseline) over the number of cases averted. The ICER is shown below as incremental cost per case averted:

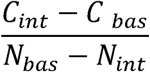

Where C_int_ and N_int_ are respectively the cost and the number of cases incurred by one of the interventions, and C_bas_ and N_bas_ are costs and cases at baseline.

Due to some interventions having ranges crossing zero, we also calculated the probabilities of these interventions being cost saving. For every run of the model, we calculated how many were cost saving compared to the baseline.

### Sensitivity analysis

To account for imperfect performance of the test, a sensitivity analysis was undertaken on the diagnostic sensitivity of the LFTs and is described in the supplementary material. For each of the 803 model runs, a random value between 68% and 76% was selected to represent the estimated sensitivity of LFTs [16]. The proportion of resident cases and deaths, as well as costs related to resident hospitalisation, staff testing, and quarantine, were then calculated.

## Results

Fig 1 displays the percentage of resident cases and deaths averted compared to the baseline scenario over a 12-month period, for each intervention. Modelling results show that testing alone (intervention 1) has limited epidemiological impact in reducing cases and deaths. During the twelve-month run time, daily staff testing alone averted up to 3.8% (95%UI: 3.1-4.5%) of resident cases and 3.5% (95%UI: 2.3-4.4%) of resident deaths compared to the baseline testing scenario. Similarly, baseline testing with a public health and social measure (intervention 2) also showed limited impact, by reducing up to 7.4% (95%UI: 5.7-9.2%) cases and 7.0% (95%UI: 5.2-9.0%) deaths compared to baseline testing alone. Contrastingly, the two interventions combined led to a decrease of more than 20% in cases and deaths when testing of staff was performed at least twice a week, with maximum impact of 53.6% (95%UI: 49.5-57.7%) cases and 49.9% (95%UI: 26.3-59.1%) deaths averted by daily testing, when compared to baseline.

**Fig 1:**
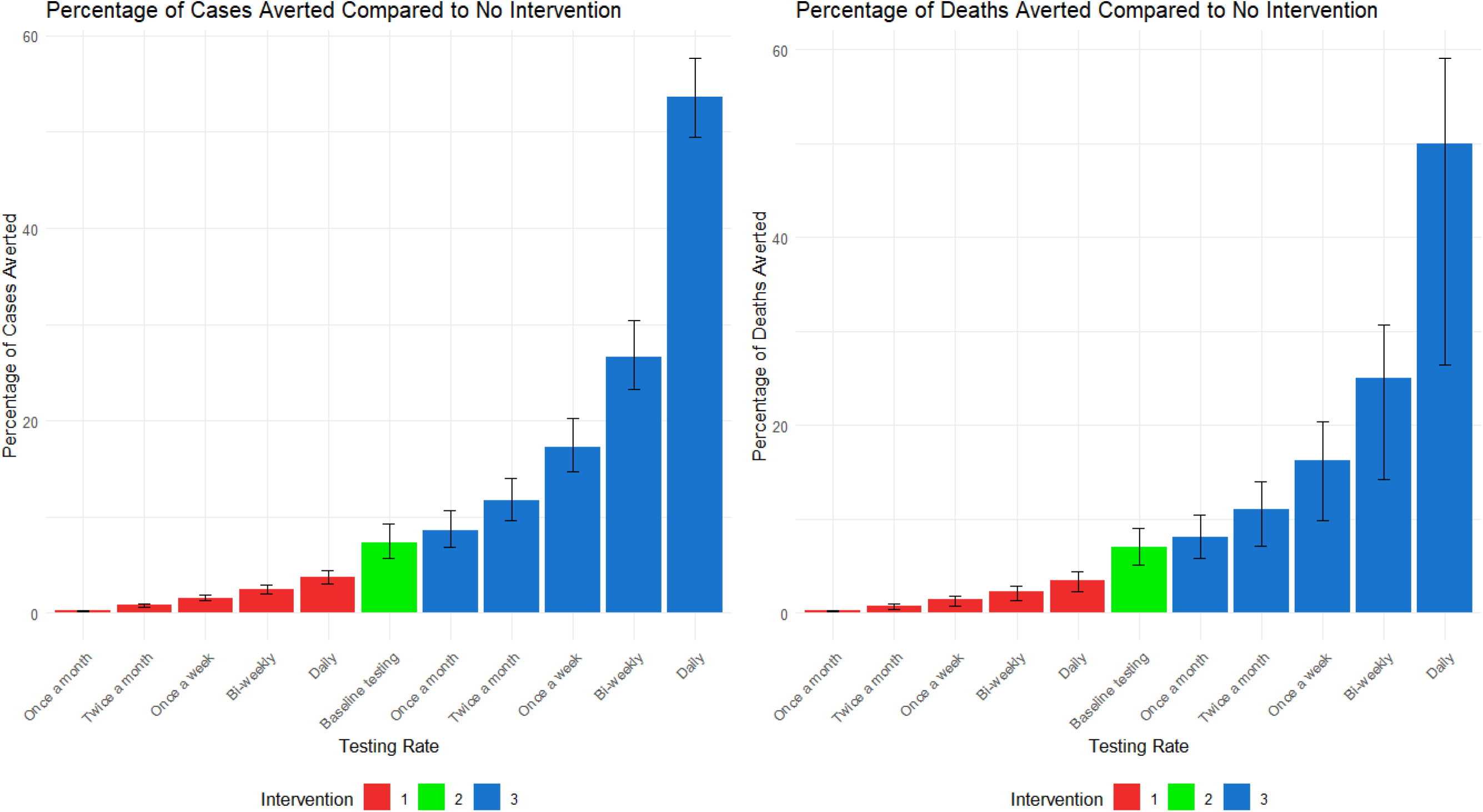
Percentage of cases and deaths averted for each intervention with 95% uncertainty interval (UI). Intervention 1: Staff are tested once a month, twice a month, once a week, twice a week and daily. Intervention 2: Baseline testing rates with a hypothetical public health and social measure that prevents the transmission of the infection between residents. Intervention 3: Staff are tested once a month, twice a month, once a week, twice a week and daily, in conjunction with the hypothetical public health and social measure. Increased staff testing alone is not enough to visibly reduce the number of cases and deaths in the resident population compared to baseline. Similarly, a public health and social measure alone has also a limited epidemiological impact. However, combining testing and public health and social measures can lead to a ∼20% reduction in cases and deaths, in the case of weekly testing, ∼30% reduction with twice-weekly testing, or up to ∼60% reduction with daily testing.

Following the epidemiological analysis, total costs were calculated for each intervention over the whole 12-month implementation period (Table 2) and associated <u>with</u> the model epidemiological results (Fig 1). The analysis shows that testing-only strategies (intervention 1) are costly, with total incremental costs ranging from £0.6 to £22 million over a 12-month period. In contrast, baseline, monthly or twice-a-month testing when combined with a hypothetical public health and social measure (interventions 2, 3a, 3b) dominate the baseline strategy, leading to significant cost savings compared to baseline testing without a public health and social measure, with savings ranging from £0.7 to £3.4 million. These savings are attributed to a reduction in resident hospitalisations and no public health and social measure related implementation costs. This means that the combined baseline/monthly/bi-monthly testing plus a public health and social measure strategy would be cost-saving only in situations where the cost of implementing the public health and social measure was no higher than £3.4 million. More frequent testing strategies, such as weekly and twice-a-week testing combined with public health and social measures, may have less substantial cost savings than less frequent testing strategies, but are cost-effective compared to baseline due to the larger number of cases averted, and have high probabilities of also being cost-saving (96.6% and 89.4%, respectively); however, this depends on the cost of the public health and social measure. In contrast, intervention 3 with daily testing is more expensive than baseline in most cases, but it may be cost-effective as it leads to strong reductions in cases and deaths. Thus, any decision on its implementation is up to policy makers depending on account of resource availability.

**Table 2:** Interventions and associated costs. Table 2 displays hospitalisation, intervention and total costs for all interventions, including baseline. Intervention costs comprise LFT tests, staff sick pay and backfill. Hospitalisation costs comprise non-critical and critical care. Intervention and hospitalisation costs are combined to give an overall cost for each intervention and are compared to the baseline. All costs are in 2022-23 millions of pounds (£).

| Intervention | Intervention costs<br>(£ million) | Hospitalisation costs<br>(£ million) | Total costs (£ million) | Costs averted (baseline<br>costs minus intervention<br>costs) (£ million) |
| --- | --- | --- | --- | --- |
| Baseline testing | 2.0 (95% UI: 1.7; 2.5) | 36.2 (95% UI: 29.9;<br>44.1) | 38.2 (95% UI: 31.6; 46.4) |  |
| Once a month testing | 2.8 (95% UI: 2.3; 3.4) | 36.1 (95% UI: 29.8; 44.0) | 38.9 (95% UI: 32.2; 47.3) | -0.7 (95% UI: -0.9; -0.6) |
| Twice a month testing | 4.5 (95% UI: 3.7; 5.5) | 35.9 (95% UI: 29.6; 43.8) | 40.4 (95% UI: 33.5; 49.0) | -2.2 (95% UI: -2.7; -1.8) |
| Once a week testing | 6.9 (95% UI: 5.8; 8.3) | 35.7 (95% UI: 29.4; 43.4) | 42.6 (95% UI: 35.3; 51.6) | -4.3 (95% UI: -5.2; -3.7) |
| Twice a week testing | 10.4 (95% UI: 9.0; 12.0) | 35.3 (95% UI: 29.1; 43.1) | 45.7 (95% UI: 38.2; 55.0) | -7.5 (95% UI: -8.6; -6.6) |
| Daily testing | 24.4 (95% UI: 22.7; 26.5) | 34.9 (95% UI: 28.7; 42.5) | 59.2 (95% UI: 51.5; 68.8) | -21.0 (95% UI: -22.4; -19.9) |
| Baseline testing with public health and social measure | 2.0 (95% UI: 1.7; 2.5) | 33.6 (95% UI: 27.5; 41.2) | 35.6 (95% UI: 29.1; 43.5) | 2.7 (95% UI: 2.0; 3.4) |
| Once a month testing with public | 2.8 (95% UI: 2.3; 3.4) | 33.1 (95% UI: 27.0; 40.7) | 35.9 (95% UI: 29.4; 44.1) | 2.3 (95% UI: 1.5; 3.1) |
| health and social<br>measure |  |  |  |  |
| Twice a month<br>testing with public<br>health and social<br>measure | 4.5 (95% UI: 3.7; 5.5) | 32.0 (95% UI: 26.0;<br>39.4) | 36.5 (95% UI: 29.9; 45.0) | 1.7 (95% UI: 0.7; 2.7) |
| Once a week<br>testing with public<br>health and social<br>measure | 6.9 (95% UI: 5.8; 8.3) | 30.0 (95% UI: 24.1;<br>37.2) | 36.9 (95% UI: 30.2; 45.6) | 1.3 (95% UI:<br>-0.07; 2.7) |
| Twice a week<br>testing with public<br>health and social<br>measure | 10.4 (95% UI: 9.0;<br>12.0) | 26.6 (95% UI: 21.2;<br>33.6) | 37.0 (95% UI: 30.4; 45.6) | 1.2 (95% UI:<br>-0.5; 2.9) |
| Daily testing with<br>public health and | 24.4 (95% UI: 22.7;<br>26.5) | 16.9 (95% UI: 12.9;<br>22.2) | 41.2 (95% UI: 35.7; 48.5) | -3.0 (95% UI:<br>-5.4; 0.3) |
| social measure |  |  |  |  |

The sensitivity analysis results showed a similar trend to that observed in Fig 1, with Intervention 3 (twice-weekly and daily staff testing) consistently averting the highest number of cases and deaths compared to lower testing frequencies and the staff-only intervention. The epidemiological impact decreased with reduced test sensitivity (up to 13% fewer cases and 27% fewer deaths averted for daily testing combined with public health and social measures). Although significant epidemiological impact was observed even at test sensitivities between 68% and 76%, these findings demonstrate the importance of using high-sensitivity tests. Test sensitivity also significantly influenced the cost-effectiveness of interventions. While three interventions were cost-saving under the assumption of perfect test sensitivity, this number decreased to two when sensitivity was reduced. Twice-weekly testing combined with public health and social measures, which appeared potentially cost-saving under perfect test sensitivity assumptions, was not cost-saving in the sensitivity analysis. These findings highlight the influence of test sensitivity on both epidemiological outcomes and economic evaluations. Further details are provided in the Supplementary Appendix.

Fig 2 presents the cost-effectiveness plane for the various intervention strategies. Interventions 2 and 3 are shown to be less costly in most instances and successful at averting resident cases. However, daily testing under intervention 3, despite averting a significant number of resident cases, may result in higher costs than the baseline scenario.

**Fig 2:**
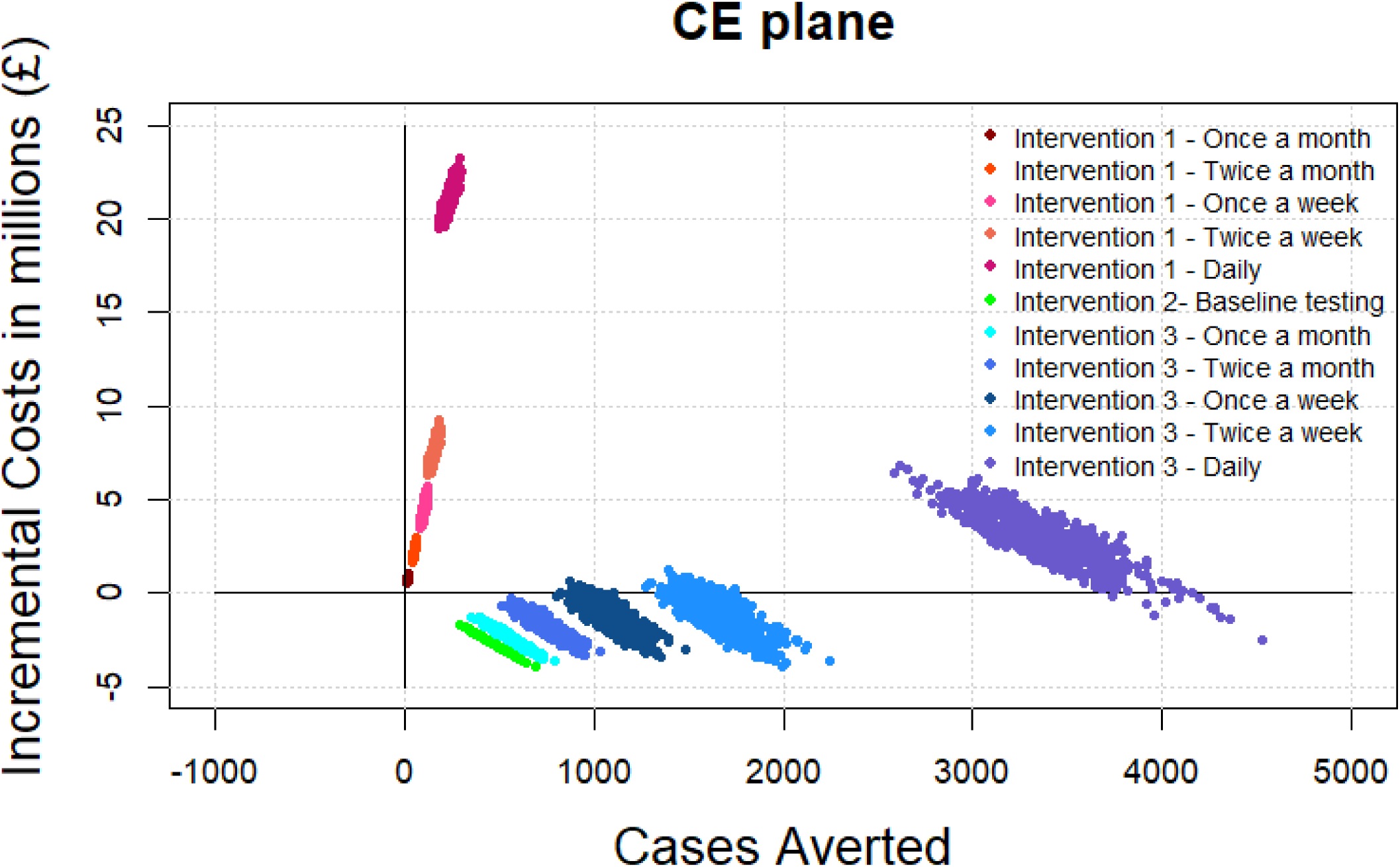
Cost-effectiveness plane as Incremental costs per COVID-19 cases averted over 12 months. Health outcomes (resident cases averted) are plotted on the X axis and incremental costs in millions (£) are plotted on the Y axis. Each point represents incremental costs over cases averted for each model run compared to the baseline scenario. Interventions 2 and 3 (with monthly, twice a month, weekly and twice a week testing) are cost-saving in most cases and always cost-effective. The cost-effectiveness of Intervention 3 with daily testing, however, depends on resource availability.

## Discussion

We presented a compartmental model simulating SARS-CoV-2 transmission in care homes for older adults in England to ascertain the impact of staff testing interventions on resident cases and deaths. We further explored the associated costs for each intervention to provide policy makers with grounded evidence to aid their decision-making for future pandemics.

The results of the study demonstrate that staff testing alone is not enough to evidence any substantial reductions in cases and deaths in the resident population. Daily testing without a public health and social measures averted only 3.8% (95%UI: 3.1-4.5%) cases and 3.5% (95%UI: 2.3-4.4%) deaths when compared to baseline. However, regular staff testing, when paired with public health and social measures such as resident zoning, using PPE, contact tracing or improved room ventilation, is the most effective strategy for reducing resident cases and deaths. Our results showed that during the early phase of a pandemic (emerging virus, no prior immunity, no vaccination) 26.6% (95%UI: 23.2-30.4%) resident cases and 25% (95%UI: 14.2-30.7%) resident deaths could be averted with twice-weekly testing of staff and 53.6% (95%UI: 49.5-57.7%) resident cases and 49.9% (95%UI: 26.3-59.1%) resident deaths with daily testing, combined with a hypothetical public health and social measure that prevents transmission among residents. Furthermore, monthly, twice-monthly, weekly and twice-weekly testing, when combined with public health and social measures, are cost-effective and most likely cost-saving interventions, and can lead to up to ∼£3.4 million costs saved compared to baseline over 12 months. These cost savings are due to a consistent reduction in hospitalisations in residents. On the other hand, daily testing combined with public health and social measures is a costly intervention, where the high cost of frequent testing appears to outweigh the financial benefits from fewer hospitalisations (up to £5.3 million additional costs compared to baseline). However, due to the high number of cases and deaths that this intervention can avert, we cannot make definitive conclusions on its cost-effectiveness, and any policy decisions should take resource availability into account.

Our epidemiological results align with several other studies, which suggest public health and social measures—such as personal protective equipment (PPE), zoning of symptomatic cases, and improved room ventilation—can be used alongside staff testing to maximise the prevention of cases and deaths[3–4]Click or tap here to enter text.. It is crucial to carefully consider the type of public health and social measure implemented in conjunction with staff testing to ensure residents’ well-being is respected. Zoning within care homes may offer a practical solution for containing infections; however, its effectiveness will depend on factors such as the facility’s layout and available staff. While our findings emphasise the importance of public health and social measures alongside staff testing in reducing infections, future guidelines must also prioritise compassion. Full resident isolation is unlikely to be the optimal approach moving forward.

Our epidemiological results also differ from studies such as McGarry et al. (2023) [16] who demonstrated that increased surveillance testing of staff members was associated with meaningful reductions in COVID-19 cases and deaths among residents-especially before vaccine availability.

When it comes to the frequency of testing, our results align with the findings of Gómez Vázquez et al. [5] who also modelled testing interventions in care homes and reported that increasing testing frequency to every three days led to a significant reduction in infections and hospitalisations. On the other Hand, Nguyen et al.[4], recommend staff testing be carried out every 7 to 10 days, explaining that although more frequent testing intervals result in improved outcomes, this may be costly and not feasible. Whilst our results do show daily testing may be more costly than the baseline scenario, we also illustrated how twice-weekly testing may be cost-saving and always cost-effective. Additionally, while more frequent testing could increase workload, time pressures and discomfort associated with testing, our study has factored in the sickness pay and backfill costs for staff required to isolate. This demonstrates that it is economically feasible to test as frequently as twice-weekly, although it is important to note that the costs of testing, backfill and sickness pay were borne by the government rather than care providers during the COVID-19 pandemic.

Our model has several strengths. This is the first study able to estimate the cost per case averted of different testing scenarios, which is important from a policy-making perspective. Additionally, our analysis is grounded in real data, having parametrised the model using actual rates of COVID-19 in care homes for older adults in England as observed during the VIVALDI study [7] in 2021-2022. At the same time, the scenario and sensitivity analyses allow us to look beyond the case of COVID19 and make our results informative for future pandemics.

Our study also has several limitations. Firstly, due to lack of available data, we could only consider a hypothetical public health and social measure and were not able to explore specific interventions aimed at reducing transmission among residents, such as partial or total resident zoning, resident testing, the use of face masks, or improved room ventilation. This affects both our epidemiological and economic results as we could not consider the costs of these interventions which should be explored in future research. We were, however, able to estimate thresholds on the maximum cost these public health and social measures should have for the combined testing plus public health and social measure strategies to be cost saving. Furthermore, due to the lack of available data, we parameterised the model with transmission rates during the beginning of the vaccination campaign-despite focusing our work on pre-vaccination. Our baseline is therefore representative of the first stages of a vaccine campaign where 38.4% of the population had a first vaccine by the 18^th^ of March [17]. This will likely underestimate the impact of interventions before vaccinations are in place. Chen et al. demonstrated that testing in care homes was highly cost-effective - 3.5 times more cost-effective prior to vaccination rollout [18].

Secondly, because our goal was to study a best-case scenario in order to estimate the maximum possible impact of testing strategies, we assumed perfect sensitivity. However, to address real-world conditions, we also conducted a sensitivity analysis (see Supplementary Appendix) using the actual sensitivity of LFT tests. This analysis demonstrated that each intervention prevents fewer cases and deaths and is less cost-effective under realistic conditions. These findings illustrate the importance of developing high-sensitivity tests to maximise epidemiological and economic outcomes.

Finally, our model was designed to represent an emergency scenario during the first stages of a new pandemic or a new viral strain of an existing infection. For this reason we made some strong assumptions: (i) we did not include visits from relatives or health professionals in our model. This may not be the case in future pandemic as more recent guidance set out by the UKHSA reinforces the importance of allowing care home residents contacts with friends and relatives [19] Our assumption is however supported by Rosello et al. [3] who estimated the importation of SARS-CoV-2 by visitors to be negligible. (ii) We also assumed that contact patterns between staff and residents remained constant throughout the simulation, but these patterns may change during an outbreak. (iii) We assumed no vaccine protection.

Assuming no protection from vaccination is reasonable in the context of care homes’ preparedness to future pandemics. However, to inform the management of COVID-19 and other infections for which vaccines are already available, it would now be beneficial to revisit these findings under varying levels of vaccine protection to assess whether testing frequencies should be adjusted in the context of vaccination. Several studies, including Fosdick et al. [20] and Vilches et al. [21] demonstrated how effective vaccines could be with low testing rates, while Kahn et al. [22] emphasised that vaccines with lower efficacy should be paired with more frequent testing, particularly in the context of waning immunity/need for re-vaccination. Exploring this further—particularly with the economic component of our analysis—could offer valuable insights. Future research could investigate different vaccine efficacies and the corresponding testing rates required, while incorporating the economic considerations that are central to policy decision-making.

## Conclusion

The study demonstrates the importance of combining staff testing with public health and social measures in order to reduce cases and deaths in care homes. Frequent asymptomatic staff testing alone has limited impact in reducing resident cases and deaths and is expensive; yet, combined with low-cost and highly effective public health and social measures, it can significantly improve epidemiological outcomes and yield substantial cost savings. Daily testing is the most effective in averting resident cases but may also result in higher costs, highlighting the balance needed between improved health outcomes and the associated financial costs. Biweekly staff testing seems to be the most effective while still being potentially cost saving. Asymptomatic staff testing, combined with public health and social measures was also shown to be superior to public health and social measures with baseline symptomatic testing, demonstrating the importance of testing. It is important to note, however, that decisions around cost effectiveness of testing may change depending on the pathogen. For instance, with a less virulent pathogen, the resulting low rates of hospital admissions may render frequent testing economically unfeasible, or vice versa, worse pathogens that resulted in higher rates of hospital admissions would make the testing interventions here explored even more cost-effective. Despite this, these findings are critical for guiding the rapid implementation of testing strategies, sick pay policies, and agency backfill measures in future public health response. Policy makers must consider the incremental costs associated with each intervention against the reduction of cases and deaths each offers, as well as the availability of resources when preparing for future epidemics and pandemics in care home settings. Furthermore, the challenges surrounding funding for staff testing, sick pay, and backfill due to increased testing and the inclusion of public health and social measures will need to be addressed, including determining whether the responsibility lies with care providers, the NHS, or the Government.

## Data Availability

All data used in this modelling study has been reported in the supplementary documentation and sources have been accordingly cited.

