## Supplementary appendix for "Staff testing in care homes for older people: policy implications for early stages of future pandemic responses"

Model schematic

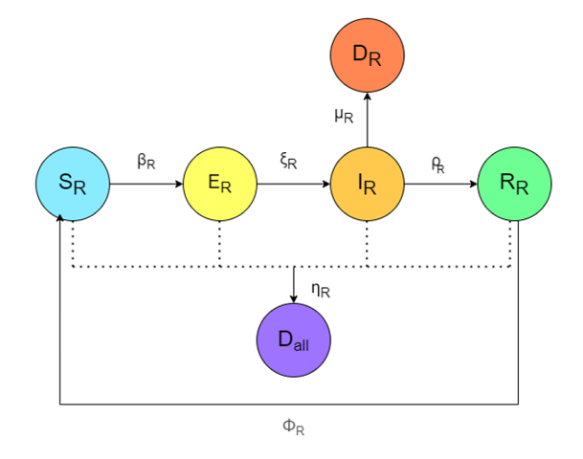

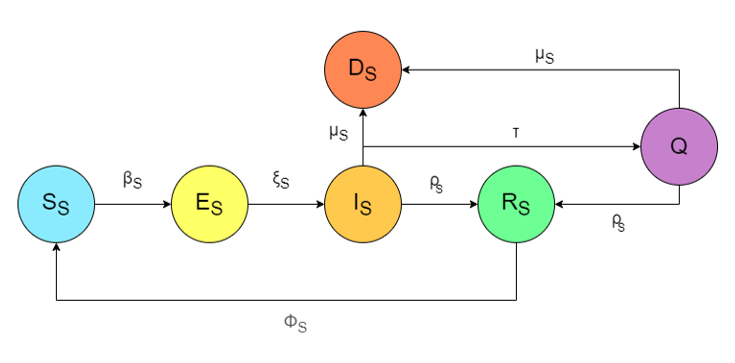

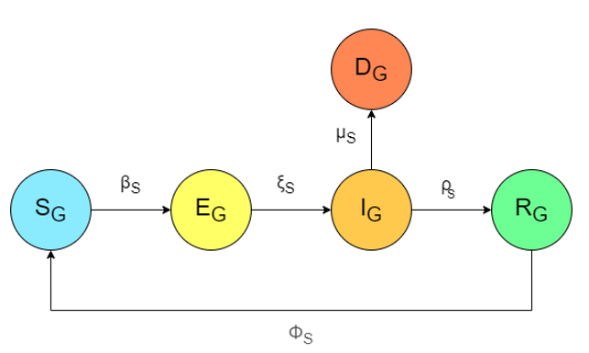

Figure 1:

SEIR model for the three population strata: Care home residents (65 years and above), staff and the general population. All rates for the general population equal that of the staff, hence the rates depicted for staff and general population in the SEIR figures differ from the residents.

Model dynamic

The model employs a susceptible-exposed-infectious-recovered (SEIR) framework. Initially, all individuals are in the susceptible compartment (S). Upon infection with SARS-CoV-2 (transmission rate β), they move to the exposed compartment (E), where they are infected but not yet capable of transmitting the virus. After an incubation period, individuals progress (at rate ξ) to the infectious compartment (I), where they can spread the virus to others. After this stage, they either recover (R), at a recovery rate (ρ), returning to the susceptible compartment after immunity wanes (rate ϕ), or they die of COVID-19 (D), at a rate of μ. Additionally, residents can die from other causes (rate η).

We chose not to model non-COVID-19 deaths among staff or the general population, as our focus was on residents, and these deaths would have minimal impact on overall population size. The SEIR framework also includes a quarantine compartment (Q), where infectious staff move after receiving a positive lateral flow test (LFT), transitioning from the infectious stage to quarantine. The whole population (N) is comprised of S+E+I+R for residents and the general population and S+E+I+Q+R for staff.

To assess the effectiveness of staff testing and non-pharmaceutical interventions (NPIs), we further adjust staff testing rates (τ) and the inclusion of an NPI (C).

Model Equations:

Residents:

**Ṡ_r_** = (η_r_ * S_r_ + η_r_ * E_r_ + η_r_ * R_r_ + μ_r_ * I_r_) + ϕ * R_r_ - λ * S_r_ - η_r_ * S_r_

**Ė_r_** = λ * S_r_- ξ_r_ * E_r_	- η_r_ * E_r_

**İ_r_** = ξ_r_ * E_r_ - (ρ_r_ + μ_r_) * I_r_

**Ṙ_r_** = ρ_r_ * _Ir_ - (ϕ + η_r_) * R_r_

**Ḋ_r_** = μ_r_ * I_r_

Staff:

**Ṡ_s_** = ϕ * R_s_ + μ_s_ * (I_s_ + Q_s_) - β_s_ * S_s_ * (I_r_ + I_s_ + I_g_) / (N_r_ + N_s_ + N_g_)

**Ė_s_** = β_s_ * S_s_ * (I_r_ + I_s_ + I_g_) / (N_r_ + N_s_ + N_g_) - ξ * E_s_

**I_̇s_** = ξ * E_s_ - (τ + ρ + μ_s_) * I_s_

**Q̇_s_** = τ * I_s_ - (ρ + μ_s_) * Q

**Ṙ_s_** = ρ * (I_s_ + Q) - ϕ * R_s_

**Ḋ_s_** = μ_s_ * I_s_ + μ_s_ * Q_s_

General Population:
**Ṡ_g_** = ϕ * R_g_ - β_g_ * S_g_ * (I_g_ + I_s_) / (N_g_ + N_s_) - (η_r_ * S_r_ + η_r_ * E_r_ + η_r_ * R_r_ + μ_g_ * I_r_)

**Ė_g_** = β_g_ * S_g_ * (I_g_ + I_s_) / (N_g_ + N_s_) - ξ * E_g_

**İ_g_** = ξ * E_g_ - (ρ + μ_g_) * I_g_

**Ṙ_g_** = ρ * I_g_ - ϕ * R_g_

**Ḋ_g_** = μ_g_ * I_g_

λ = [ β_r_ * (c*I_r_ + I_s_) / (N_r_ + N_s_)]

c is a factor representing the inclusion of the NPI. The NPI reduces transmission among residents; in our analysis we set c = 0 when the NPI is considered, and c = 1 when not.

Table 1: Parameter Table

Table 1 shows the parameters included in the model, and the prior and posterior values. ‘Parameter’_r represents values for residents, otherwise values displayed are for the general population and staff. Numbers following this illustrate how these are time varying parameters

| **Parameter** | **Meaning** | **Prior Value** | **Posterior Value** | **Reference** |
| --- | --- | --- | --- | --- |
| ϕ | Loss of immunity | 0.1-0.2 | 0.11 - 0.19 | [1] |
| β | Transmission rate |  | β_r1_ = 5.40 - 8.42  β_r2_ = 12.40 - 22.40  β_r3_ = 30.05 - 95.90  β_r4_ = 93  β_1_ = 0.27 - 2.31  β_2_ = 3.00 - 3.85  β_3_ = 4.36 - 5.64  β_4_ = 6.03 - 9.81  β_5_ = 11.25 | Calibration |
| ξ | Incubation period | ξ_r_ = 3.57-7.35  ξ = 3.57-8.47 | ξ_r_ = 3.83 - 7.14  ξ= 3.79 - 8.13 | [2-3] |
| ρ | Recovery rate | ρ_r_ = (2.3,3.1)  ρ = (2.41,3.18) | ρ_r_ = 2.42 - 3.06  ρ = 2.56 - 3.13 | [4-5] |
| μ | COVID-19 death rate | μ_r1_ = (0.0031, 0.31)  μ_r2_ = (0.00031, 0.28)  μ_1_ = (0.0031,0.031),  μ_2_ = (0.00031,0.0031) | μ_r1_ = 0.02 - 0.30  μ_r2_ = 0.02 - 0.27  μ_r3_ = 0.02 - 0.30  μ_1_ = 0.01 - 0.02  μ_2_ = 0.0005 - 0.0030  μ_3_ = 0.01 - 0.02 | [6-7] , including calibration |
| η | All causes mortality | 0.01-0.03 | 1.074896e-02 - 2.825495e-02 | [7] |
| τ | Testing rate | τ = 0.54-0.85 | τ = 0.59-0.73 | Calibration [8] |

Transmission

We account for differences in transmission between residents, staff, and the general population by assigning distinct transmission values to each group, denoted as β_r_ for residents and β for staff and the general population. This is to account for the different mixing each group has. Additionally, we adjust these transmission values for specific months to reflect changes due to lockdown measures ([8], emerging strains [10-12] , and vaccination rates [13].

β_r1_ and β_1_ account for the transmissibility of the strain in January 2021. Early 2021, B.1.1.7 (Alpha Variant) became dominant [13]; hence we used this as our reference point [15]. Lockdown measures legally came into force on the 26th of March and a conditional plan to lift lockdown was announced on the 10th of May [9]. We introduced β_2_ into the model to reflect this period of lockdown which affects primarily the general population and staff. β_3_ denotes the strain in November 2021, where BA.5 (an Omicron variant) was dominant [16], as well as the introduction of the second national lockdown on the 5^th^ of November [9]. Omicron was more infectious than the previous strains [10] so we account for this through β_3_ and β_r2._ β_4_ and β_r3_ refer to February and March 2022, with BA.1 and BA.2 (Omicron Variants) driving high numbers of infections [17], and accounts for the third national lockdown [9].

The scenario analysis used a transmission parameter that is 1.15-1.87 times higher than the previous value to account for a new strain with increased infectiousness. We also changed the mortality values to equal the values witnessed at the beginning of the pandemic to reflect a more deadly virus.

Testing

For each intervention the testing rate (τ) varied. For the baseline scenario, staff members were tested less than once a month (~0.6). This value was estimated from the VIVALDI study [7] and reflects staff members who only test when symptomatic. For each intervention, we varied the testing rate to equal 1,2,4,8,30 which reflects once-a-month, twice-a-month, once-a-week, twice-a-week and daily staff testing.

Calibration targets

Table 2: Calibration Targets Table

| Target name | Month | Target Values | Reference |
| --- | --- | --- | --- |
| COVID-19 cases in Residents in England | September 2021 | 85-187 | [4] |
|  | October 2021 | 161-360 |  |
|  | January 2022 | 916-1,253 |  |
|  | March 2022 | 704-1,050 |  |
| COVID-19 cases in the general population in England | September 2021 | 319,086-699,750 | [3] |
|  | October 2021 | 733,338-1,220,364 |  |
|  | January 2022 | 1,640,214-3,834,630 |  |
|  | March 2022 | 2,955,744-4,551,174 |  |
| Total COVID-19 deaths | January 2022 | 50859-76289 | [18] |

Calibration Description

The model was developed using R (v4.3.3) and calibrated with the History Matching and Emulation (hmer) package, which utilises Bayes Linear methods for emulation and history matching. Calibration was based on observed COVID-19 prevalence data from the general population and care home staff and residents, using data from the VIVALDI study (ISRCTN14447421) between January 2021 and March 2022. Calibration targets are reported in Table 2.

To ensure efficient exploration of the parameter space, we built emulators that are computationally fast and capable of generating predictions in unexplored regions. These emulators incorporate statistical measures to capture uncertainty at any point in the parameter space. The emulation process was integrated with history matching, a technique that systematically eliminates implausible parameter regions, narrowing down the range of parameter values that are consistent with observed data. From this calibration, 803 parameter sets were generated and used to run the model across each set.

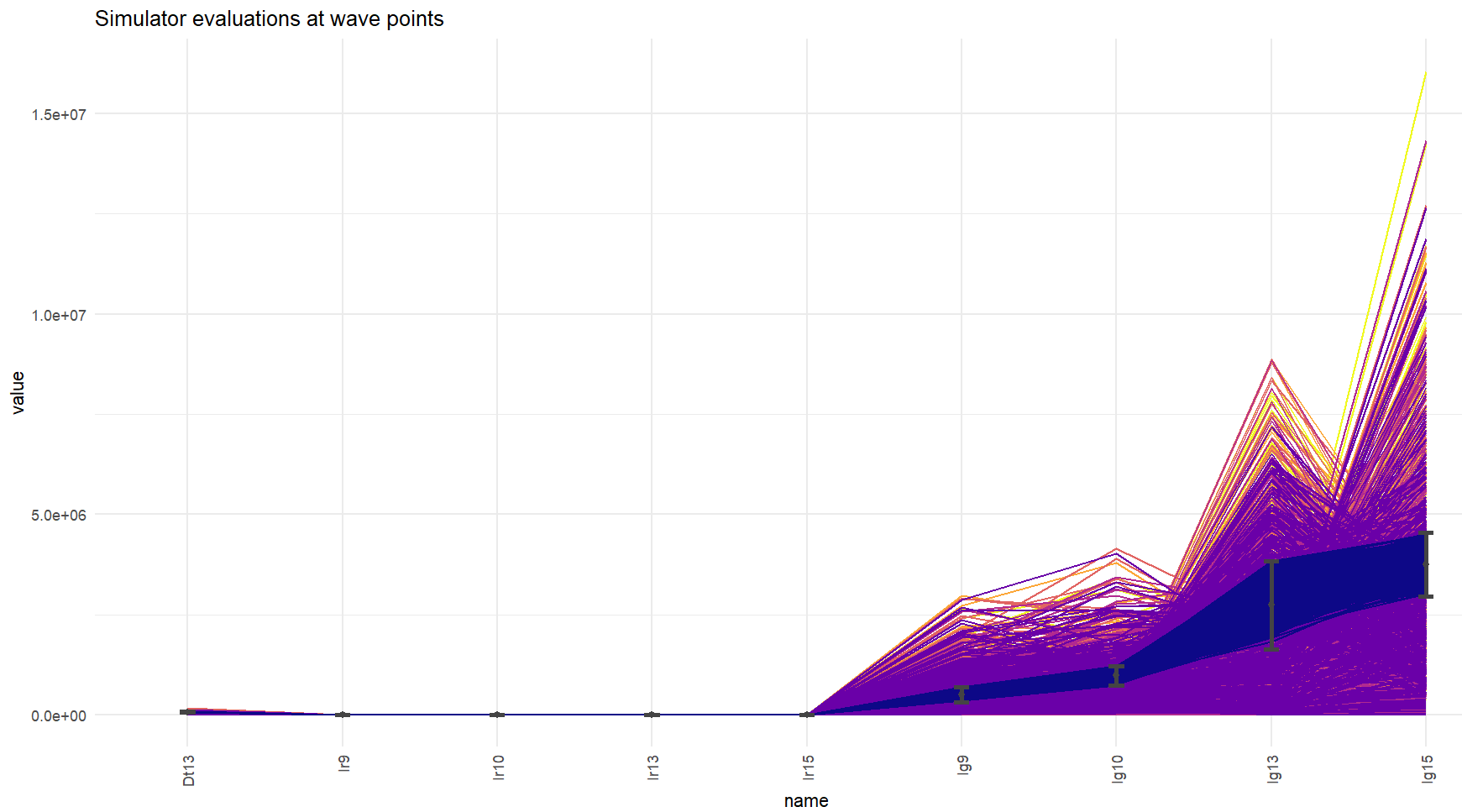

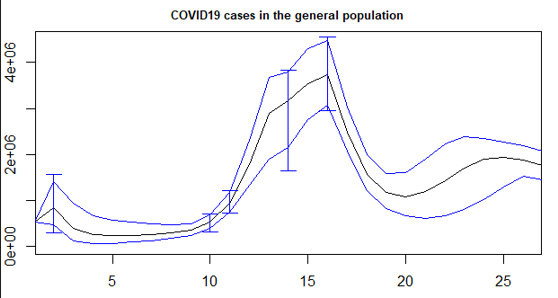

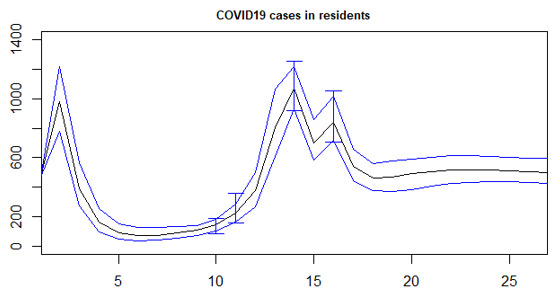

**Figure 2:** Calibration plots.

Cost Data

#### Intervention Costs

The average cost per intervention for a single staff member was calculated, incorporating expenses for asymptomatic testing, sick pay, and staff backfill. The unit costs for LFT (lateral flow tests) as specified in the contract between NIHR and UK Health Security Agency (UKHSA). Costs related to sick pay and agency staff backfill were derived from trial data collected by the UCL CCTU.

Key assumptions for intervention costs included:

- Unit costs were based on day rates from UKHSA.
- All testing occurred at work, assuming no other duties were performed during the test.
- The average testing time was assumed to be 10 minutes.
- Sick pay and backfill costs were calculated per episode, only for shifts eligible for reimbursement by UKHSA, following a positive test.

The audit data was drawn from a sample of 24 care homes, representing 180 staff members with COVID-related sickness.

#### Breakdown of Costs:

- LFT Tests: The total cost for asymptomatic testing (2x/week) per staff member per home per week, weighted by the proportion of staff participating in testing, was £7.16.
- Staff Sickness Pay and Backfill:
  - Sick Pay and Backfill Costs per Staff Episode (Calculated from audit data provided by the Trials Unit):
    - Sick Pay: £372.26
    - Agency Backfill: £155.36
    - Total Sick Pay and Backfill: £527.62

#### Hospitalisation Costs

Hospitalisation costs were calculated for each intervention, separating the costs into critical care and non-critical care. Resident hospitalisations were estimated using model outputs, based on the proportion of residents infected with SARS-CoV-2 in each scenario. This was further refined by the percentage of cases requiring either critical or non-critical care, as outlined in a report by Ferguson et al. (2020) on COVID-19 severity by age group during the first wave in England. The costs for critical and non-critical care were sourced from the National Institute for Health and Care Excellence (NICE) (2023).

For non-ICU hospitalisations we used the ordinal scale 4 from the NICE estimates, with a single non-ICU hospitalisation costing £16,159.77 and for hospitalisations requiring ICU we used the ordinal scale 5 with a single hospitalisation ICU case costing £24,809.70. These costs comprise the general ward unit cost per finished consultancy episode (FCE), the number of FCEs per admission, duration of stay in the general ward and the general ward cost per day. We assumed ICU hospitalisations included individuals requiring any form of supplemental oxygen (such as low-flow oxygen), while non-ICU cases did not require supplemental oxygen but did need ongoing medical care. Costs were considered consistent across all non-ICU hospitalisations, with the same approach applied to ICU hospitalisations.

*Sensitivity analysis*

To account for imperfect test sensitivity, a sensitivity analysis was conducted. For each of the 803 model runs, a random value between 68% and 76% was selected to represent the estimated sensitivity of LFTs [19] . The proportion of resident cases and deaths, as well as costs related to resident hospitalisation, staff testing, and quarantine, were then calculated.

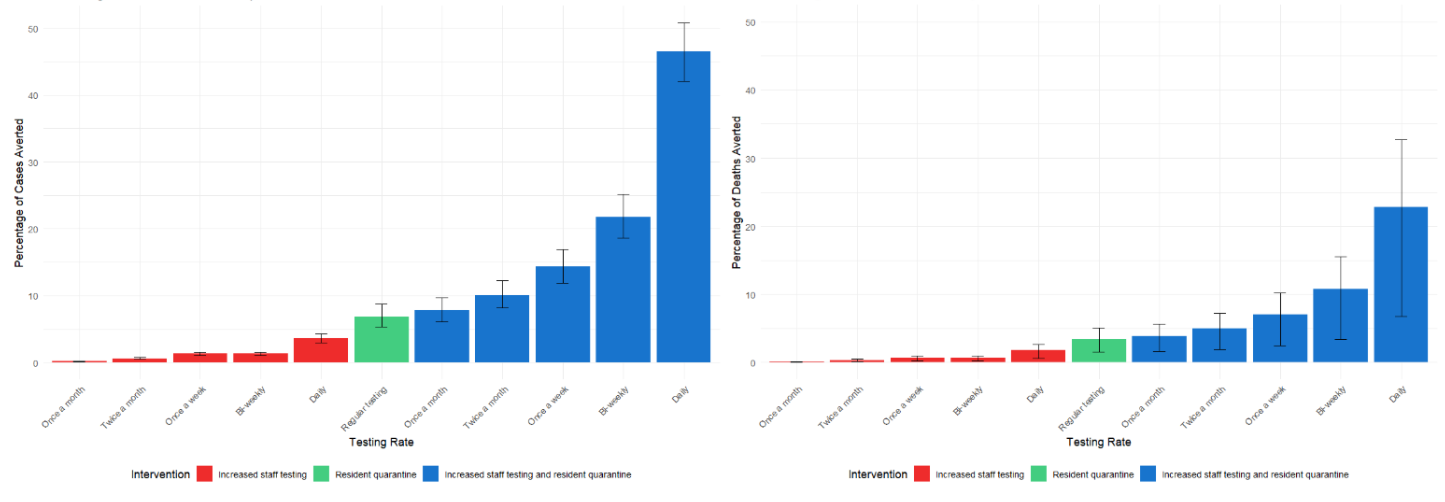

**Figure 3**: Percentage of cases and deaths averted for each intervention with 95% uncertainty interval (UI).
Intervention 1: Staff are tested once a month, twice a month, once a week, twice a week and daily. Intervention 2: Baseline testing rates with a hypothetical public health and social measure that prevents the transmission of the infection between residents. Intervention 3: Staff are tested once a month, twice a month, once a week, twice a week and daily, in conjunction with the hypothetical public health and social measure. Reduced cases and deaths averted are shown compared to 100% sensitive LFT tests.

**Table 3: Interventions and associated costs**

Table x displays hospitalisation, intervention and total costs for all interventions, including baseline. Intervention costs comprise LFT tests, staff sick pay and backfill. Hospitalisation costs comprise non-critical and critical care. Intervention and hospitalisation costs are combined to give an overall cost for each intervention and are compared to the baseline. All costs are in 2022-23 millions of pounds (£).

| **Intervention** | **Intervention costs (£ million)** | **Hospitalisation costs (£ million)** | **Total costs (£ million)** | **Costs averted (baseline costs minus intervention costs) (£ million)** |
| --- | --- | --- | --- | --- |
| Baseline testing | 2.1 (95% UI: 1.7; 2.6) | 36.3 (95% UI: 29.9; 44.2) | 38.4 (95% UI: 31.7; 46.6) |  |
| Once a month testing | 3.0 (95% UI: 2.4; 3.6) | 36.2 (95% UI: 29.8; 44.1) | 39.2 (95% UI: 32.4; 47.5) | -0.8 (95% UI:  -0.7; -1) |
| Twice a month testing | 4.9 (95% UI: 4.0; 6.0) | 36.0 (95% UI: 29.7; 43.9) | 40.9 (95% UI: 33.8; 49.8) | -2.6 (95% UI:  -2.1; -3.2) |
| Once a week testing | 7.7 (95% UI: 6.4; 9.3) | 35.8 (95% UI: 29.5; 43.6) | 43.5 (95% UI: 36.1; 52.9) | -5.2 (95% UI:  -4.3; -6.3) |
| Twice a week testing | 11.7 (95% UI: 10.0; 13.9) | 35.5 (95% UI: 29.2; 43.3) | 47.2 (95% UI: 39.5; 57.3) | -8.9 (95% UI:  -7.7; -10.5) |
| Daily testing | 26.6 (95% UI: 24.4; 29.5) | 35.0 (95% UI: 28.8; 42.6) | 61.6 (95% UI: 53.4; 72.4) | -23.3 (95% UI:  -21.6;  -25.6) |
| Baseline testing combined with public health and social measures | 2.1 (95% UI: 1.7; 2.6) | 33.8 (95% UI: 27.7; 41.5) | 35.9 (95% UI: 29.4; 43.9) | 2.5 (95% UI: 1.8; 3.2) |
| Once a month testing combined with public health and social measures | 3.0 (95% UI: 2.4; 3.6) | 33.5 (95% UI: 27.3; 41.1) | 36.4 (95% UI: 29.8; 44.7) | 1.9 (95% UI: 1.2; 2.7) |
| Twice a month testing combined with public health and social measures | 4.9 (95% UI: 4.0; 6.0) | 32.6 (95% UI: 26.6; 40.2) | 37.5 (95% UI: 30.8; 46.3) | 0.8 (95% UI: -0.3; 1.9) |
| Once a week testing combined with public health and social measures | 7.7 (95% UI: 6.4; 9.3) | 31.1 (95% UI: 25.1; 38.5) | 38.8 (95% UI: 31.8; 48.1) | -0.5 (95% UI:  -2.1; 1) |
| Twice a week testing combined with public health and social measures | 11.7 (95% UI: 10.0; 13.9) | 28.4 (95% UI: 22.7; 35.7) | 40.2 (95% UI: 33.0; 49.7) | -1.8 (95% UI:  -0.1; -3.9) |
| Daily testing combined with public health and social measures | 26.6 (95% UI: 24.4; 29.5) | 19.5 (95% UI: 15.0; 25.8) | 46.1 (95% UI: 39.6; 55.1) | -7.8 (95% UI:  -5; -10.5) |

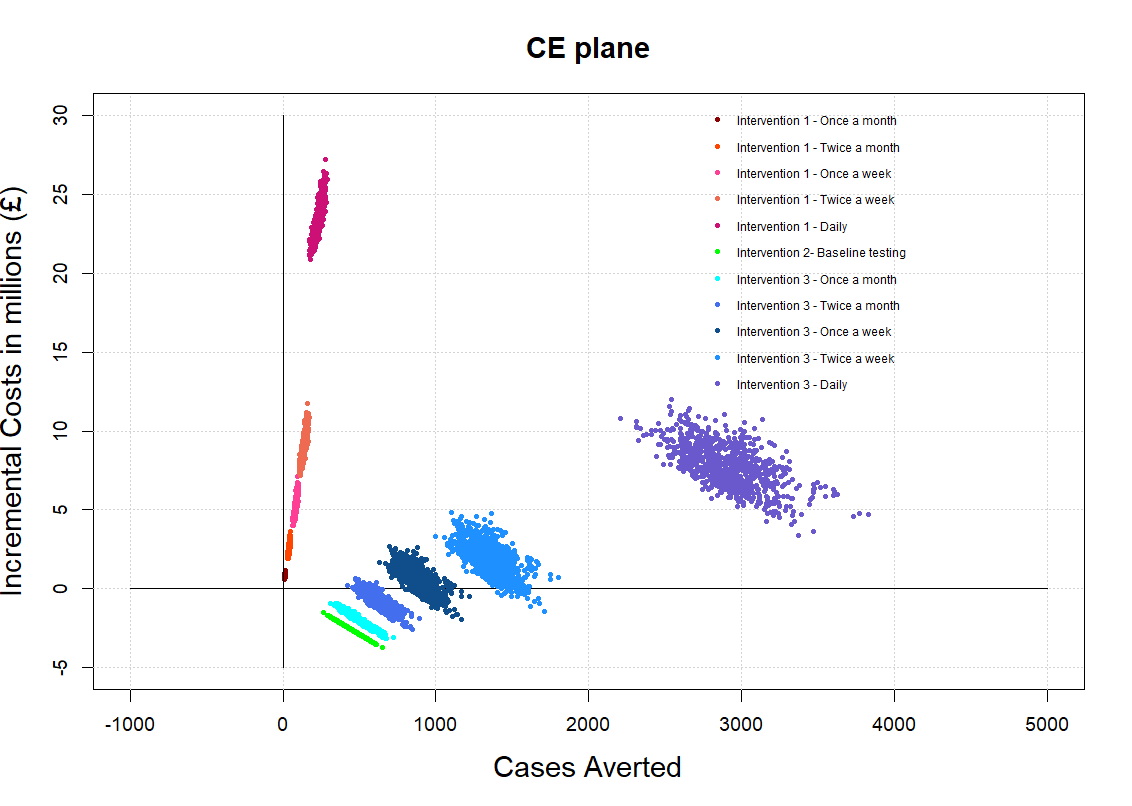

**Figure 4**: Cost-effectiveness plane as Incremental costs per COVID-19 cases averted over 12 months. Health outcomes (resident cases averted) are plotted on the X axis and incremental costs in millions (£) are plotted on the Y axis. Each point represents incremental costs over cases averted for each model run compared to the baseline scenario. When accounting for lower sensitivity, fewer interventions become cost effective.

1.	Özüdoğru O, Bahçe YG, Acer Ö. SARS CoV-2 reinfection rate is higher in the Omicron variant than in the Alpha and Delta variants. Irish Journal of Medical Science (1971 -). 2023 Apr 17;192(2):751–6.

2.	Cortés Martínez J, Pak D, Abelenda-Alonso G, Langohr K, Ning J, Rombauts A, et al. SARS-Cov-2 incubation period according to vaccination status during the fifth COVID-19 wave in a tertiary-care center in Spain: a cohort study. BMC Infect Dis. 2022 Nov 9;22(1):828.

3.	Galmiche S, Cortier T, Charmet T, Schaeffer L, Chény O, von Platen C, et al. SARS-CoV-2 incubation period across variants of concern, individual factors, and circumstances of infection in France: a case series analysis from the ComCor study. Lancet Microbe. 2023 Jun;4(6):e409–17.

4.	Kissler SM, Fauver JR, Mack C, Tai CG, Breban MI, Watkins AE, et al. Viral Dynamics of SARS-CoV-2 Variants in Vaccinated and Unvaccinated Persons. New England Journal of Medicine. 2021 Dec 23;385(26):2489–91.

5.	Menni C, VAM, PL, AM, PS, NA, . . . STD. Symptom prevalence, duration, and risk of hospital admission in individuals infected with SARS-CoV-2 during periods of omicron and delta variant dominance: a prospective observational study from the ZOE COVID Study. 2022;

6.	Nyberg T, Ferguson NM, Nash SG, Webster HH, Flaxman S, Andrews N, et al. Comparative analysis of the risks of hospitalisation and death associated with SARS-CoV-2 omicron (B.1.1.529) and delta (B.1.617.2) variants in England: a cohort study. The Lancet. 2022 Apr;399(10332):1303–12.

7.	Nomis - Official Census and Labour Market Statistics. Official census and labour market statistics.

8.	Krutikov M, Bone D, Stirrup O, Bruton R, Azmi B, Fuller C, et al. VIVALDI Cohort Profile: Using linked, routinely collected data and longitudinal blood sampling to characterise COVID-19 infections, vaccinations, and related outcomes in care home staff and residents in England. Wellcome Open Res. 2024 Aug 19;8:553.

9.	Institute for Government. Timeline of UK government coronavirus lockdowns and restrictions. 2022.

10.	UK Health Security Agency. COVID-19 variants: genomically confirmed case numbers. 2022.

11.	Duong B V., Larpruenrudee P, Fang T, Hossain SI, Saha SC, Gu Y, et al. Is the SARS CoV-2 Omicron Variant Deadlier and More Transmissible Than Delta Variant? Int J Environ Res Public Health. 2022 Apr 11;19(8):4586.

12.	Mahase E. Covid-19: How many variants are there, and what do we know about them?: Video 1. BMJ. 2021 Aug 19;n1971.

13.	Office for National Statistics. Coronavirus (COVID-19) latest insights: Vaccines. 2023.

14.	Challen R, Brooks-Pollock E, Read JM, Dyson L, Tsaneva-Atanasova K, Danon L. Risk of mortality in patients infected with SARS-CoV-2 variant of concern 202012/1: matched cohort study. BMJ. 2021 Mar 9;n579.

15.	Michaelsen TY, Bennedbæk M, Christiansen LE, Jørgensen MSF, Møller CH, Sørensen EA, et al. Introduction and transmission of SARS-CoV-2 lineage B.1.1.7, Alpha variant, in Denmark. Genome Med. 2022 Dec 4;14(1):47.

16.	UK Health Security Agency. COVID-19 variants identified in the UK – latest updates. 2023.

17.	Scientific Advisory Group for Emergencies. SAGE 105 minutes: Coronavirus (COVID-19) response, 10 February 2022. 2022.

18.	Office for National Statistics. Coronavirus (COVID-19). 2022.

19.	Krishnamoorthy A, Chandrapalan S, JalayeriNia G, Hussain Y, Bannaga A, Lei II, et al. Influence of seasonal and operator variations on diagnostic accuracy of lateral flow devices during the COVID-19 pandemic: a systematic review and meta-analysis. Clinical Medicine. 2023 Mar;23(2):144–50.
